# Estimation of stroke outcomes based on time to thrombolysis and thrombectomy

**DOI:** 10.1101/2020.07.18.20156653

**Authors:** Michael Allen, Kerry Pearn, Ken Stein, Martin James

## Abstract

**Background & Motivation:** Stroke outcomes following revascularization therapy (intravenous thrombolysis, IVT, and/or mechanical thrombectomy, MT) depend critically on time from stroke onset to treatment. Different service configurations may prioritise time to IVT or time to MT. In order to evaluate alternative acute stroke care configurations, it is necessary to estimate clinical outcomes given differing times to IVT and MT.

**Method:** Model using an algorithm coded in Python. This is available at https://github.com/MichaelAllen1966/stroke_outcome_algorithm

**Results:** We demonstrate how the code may be used to estimate clinical outcomes given varying times to IVT and MT.

**Conclusion:** Python code has been developed and shared to enable estimation of clinical outcome given times to IVT and MT. Here we share pseudocode and links to full Python code.

## 1. Introduction

In England, Wales and Northern Ireland, 85,000 people each year are hospitalised with stroke[1]. Standard care for eligible patients with acute ischaemic stroke within 4.5 hours of onset is use of intravenous thrombolysis (IVT) with alteplase, a recombinant form of tissue plasminogen activator (tPA)[2]. Time from onset to treatment is especially critical, with the effectiveness of IVT declining rapidly in the first few hours[3]. In England the proportion of stroke patients receiving IVT is about 11%[1], although higher rates have been achieved as a result of reconfiguration of urban hyperacute services[4], with maximum rates of about 20%[1].

Of patients with acute ischaemic stroke, approximately 40% have a large vessel occlusion (LVO)[5]. More recently mechanical thrombectomy (MT) has shown substantially improved clinical outcomes for patients with LVO[6]. MT is effective up to 6 hours or more after stroke onset, depending on patient selection, but also demonstrates steeply reducing benefit with increasing time from stroke onset[7,8]. The proportion of all stroke patients eligible for MT within 6 hours of onset in the UK has been estimated at about 10%[9].

Providing MT presents a significant challenge for health services. The procedure is typically carried out by a neuro-interventionist and requires support from a theatre team (anaesthetist, nurses, radiographers) and additional imaging work up, usually with computed tomography angiography (CTA), sometimes in association with other advanced imaging techniques (CT Perfusion or MRI). These additional staffing/infrastructure demands for MT require a more centralised model of service provision than currently employed for IVT[10].

Two models of hyperacute stroke care have been described and compared[11]. In a mothership model all services are provided by combined IVT/MT units, at the expense of greater travel times for some patients. In a drip and ship model, IVT may be delivered at local IVT-only units before transfer of an eligible subset of patients with LVO to a combined IVT/MT unit. Currently, neither model has been decisively shown to be superior in terms of survival or a favourable functional outcome[11], particularly given the inherent biases in observational studies[12]. The choice between these models therefore generally depends on geography and travel times, availability of experienced staff, urban/rural split, and other factors, including the maximum practical size of a combined IVT/MT unit under a mothership configuration, and the minimum required size of a IVT-only unit in a drip and ship model. The advantages and disadvantages of various system configurations may be usefully clarified by computer modelling[13,14].

The modified Rankin Scale (mRS)[15] is a commonly used scale for measuring the degree of disability or dependence in the daily activities of people who have suffered a stroke. It has been used as the primary endpoint in trials of both IVT and MT. This provides a scale of 0-6, from no disability to death (table 1).

**Table 1:** Description of modified Rankin Scale (mRS) level.

| mRS | Description |
| --- | --- |
| 0 | No symptoms at all |
| 1 | No significant disability despite symptoms; able to carry out all usual duties and activities |
| 2 | Slight disability; unable to carry out all previous activities, but able to look after own affairs without assistance |
| 3 | Moderate disability; requiring some help, but able to walk without assistance |
| 4 | Moderately severe disability; unable to walk without assistance and unable to attend to own bodily needs without assistance |
| 5 | Severe disability; bedridden, incontinent and requiring constant nursing care and attention |
| 6 | Dead |

In this paper we present a method, pseudocode, and link to python code on calculating the probability of a good outcome (mRS 0-1 at 3-6 months) in stroke, given thrombolysis and/or thrombectomy, with outcome depending on times from stroke onset to treatment. The method is based on that previously published by Holodinsky *et al*.[13].

## 2. Calculations and pseudocode

The code, and a Jupyter Notebook demonstrating use, is available at: https://github.com/MichaelAllen1966/stroke_outcome_model

DOI: 10.5281/zenodo.3947947

Estimation of clinical outcome uses a modification of a method described by Holodinsky et *al*.[13]. The model calculates the probability of a *good outcome*. A *good outcome* is taken as a patient with a mRS of 0-1 at 3-6 months (this is effectively disability-free, and may also be considered an excellent clinical outcome)[9].

The code below is presented as pseudocode and describes all the essential steps used in calculating clinical outcome (the actual code is adjusted to make use of NumPy arrays for faster calculation).

The decay in effect of IVT and ET is shown in figure 1, and comes from metaanalysis of clinical trial data for IVT[3] and MT[8].

**Figure 1.**
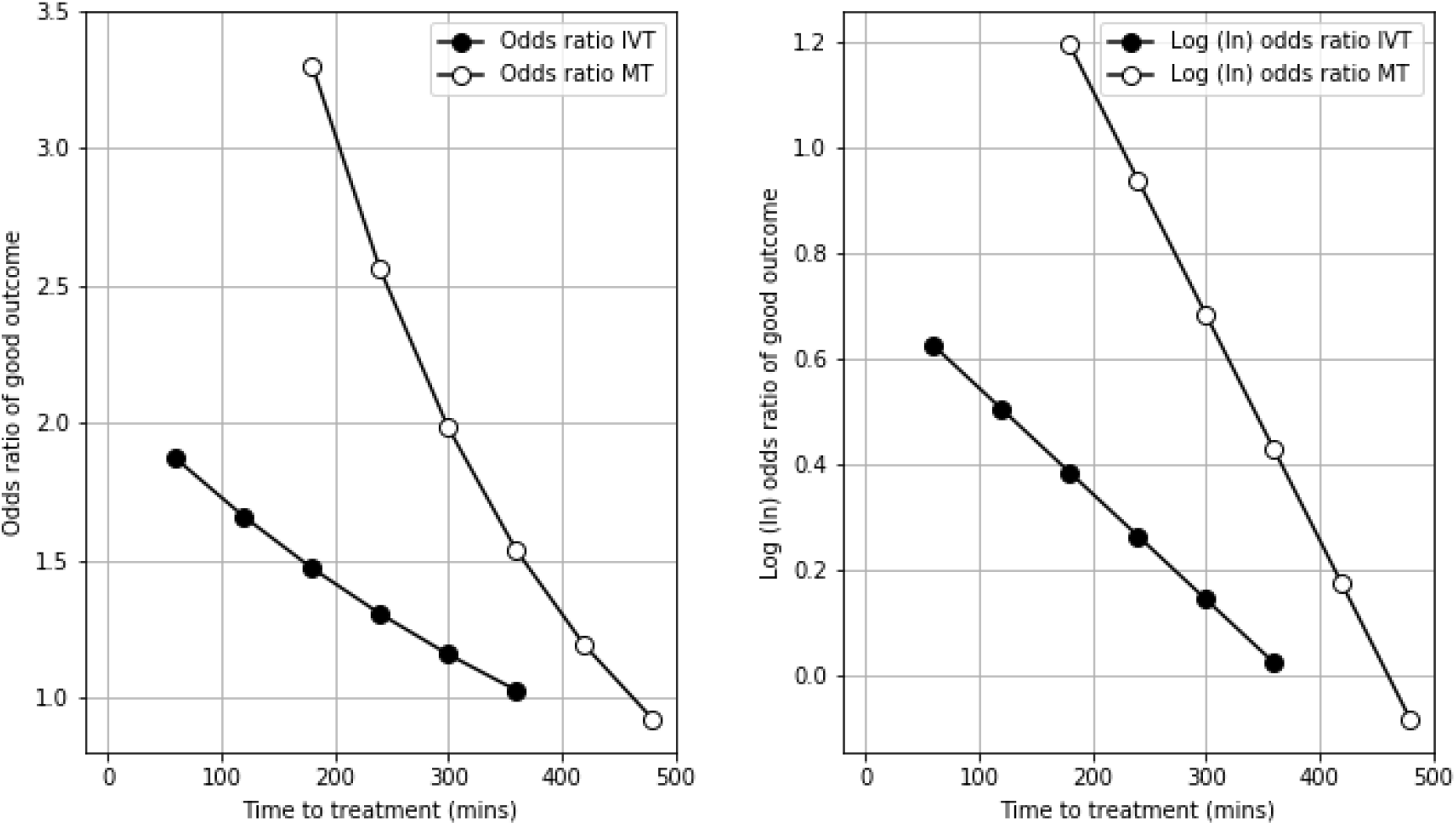
The relationship between time to treatment and odds ratio (left panel) and log (ln) odds ratio (right panel) for good (mRS 0-1) outcome after treatment with IVT (closed circles) or MT (open circles).

### 2.1. Inputs/output

All inputs take NumPy arrays (for multiple groups of patients).

`proportion_mimic`: proportion of patients with stroke mimic

`proportion_ICH`: proportion of patients with intracerebral haemorrhage (ICH). Or probability of a patient having an ICH, when using for a single patient.

`proportion_nLVO`: proportion of patients with non-large vessel occlusions (nLVO). Or probability of a patient having an nLVO, when using for a single patient.

`proportion_LVO`: proportion of patients with large vessel occlusions (LVO). Or probability of a patient having a LVO, when using for a single patient.

`onset_to_needle`: minutes from onset to thrombolysis

`onset_to_puncture`: minutes from onset to thrombectomy

`nLVO_eligible_for_treatment`: proportion of patients with nLVO suitable for treatment with thrombolysis. Or probability of a patient with NVLO being eligible for treatment.

`LVO_eligible_for_IVT`: proportion of patients with LVO suitable for treatment with thrombolysis with or without thrombectomy. Or probability of a patient with LVO being eligible for IVT.

`LVO_IVT_eligible_MT`: proportion of patients with LVO patients who are eligible to receive MT after IVT. Note: Some patients may receive MT but not IVT. This may be included here by inflating this proportion (e.g. to greater than 1.0).

The algorithm returns:

`overall_probability_good_outcomes`: The probability of having a good outcome (mRS 0-1) for the patient or group of patients (NumPy array).

### 2.2. Stroke types and mimics

In our base case we use the breakdown of stroke types as described by de la Ossa *et al*. [5]:

~~~
proportion_mimic = 0.0
proportion_ICH = 0.166
proportion_nLVO = 0.517
proportion_LVO = 0.317
~~~

*The above breakdown does not include stroke mimics. ‘Stroke mimic’ is not a clearly defined term, as whether a patient is considered a mimic or not will depend on the skill of the assessor, and the diagnostic tools available to them. However, systematic reviews of the proportion of patients with suspected stroke who had not actually had a stroke (a stroke mimic) have concluded that overall stroke mimics accounted for a quarter to a third of suspected stroke patients, depending on where in the pathway the patient is defined as a mimic[16–18]. For every 100 stroke admissions, mimics would add about another 35 arrivals or admissions. When calculating outcomes, if we include mimics, we assume mimics have a good outcome, in that they do not have a stroke-related bad outcome.

#### 2.2.1. Proportion of patients eligible for treatment

We have taken a pragmatic approach to the estimation of the proportion of patients eligible for treatment. For LVO, McMeekin *et al*.[9] have estimated that 10% of the total emergency stroke admissions may be suitable for MT. For IVT we note from SSNAP that there appears to be an upper bound of proportion of patients who receive IVT, and this occurs at 20%. Back-calculating from those figures, and the assumed breakdown of stroke types, produces the following proportions of patients suitable for treatment:

~~~
nLVO_eligible_for_treatment = 0.194
LVO_eligible_for_treatment = 0.313
~~~

In a base-case model we assume that all LVO patients who receive IVT are eligible for MT if they do not respond to IVT. In the same base-case model we assume no-one MT without first receiving IVT.

~~~
LVO_IVT_eligible_MT = 1.0
~~~

No patients with ICH, are are a stroke mimic, are eligible for treatment with IVT or MT.

### 2.3. Baseline (untreated) good outcomes

The baseline good outcomes are the proportion of untreated patients who have a good outcome (mRS 0-1)

~~~
p_good_mimic = 1.0
p_good_ICH = 0.24
p_good_nLVO_untreated = 0.462
p_good_LVO_untreated = 0.133
~~~

The untreated good outcomes for nLVO and LVO come from the placebo controlled meta-analysis of the effectiveness of IVT[3]. The severity of stroke systems (using the NIH Stroke Scale, NIHSS) is taken as an indicator of the presence of a LVO. An NIHSS of 0-10 is taken as a surrogate for a nLVO, and a NIHSS of 11 or more is taken as a surrogate for a LVO.

When calculating outcomes, if we include mimics, we assume mimics have a good outcome, in that they do not have a stroke-related bad outcome.

As patients with ICH are not eligible for IVT or MT, good outcomes for ICH are always set at their baseline untreated good outcome level.

### 2.4. Calculate additional good outcomes for patients receiving treatment

#### 2.4.1. Calculate additional good outcomes for nLVO (for those eligible for treatment)

Additional good outcomes for nLVO are based on treatment with IVT only. Additional good outcomes are good outcomes due to treatment (above the proportion of good outcomes of untreated patients).

The maximum effectiveness of IVT (alteplase), and the decay of effect of IVT was taken from the meta-analysis by Emberson *et al*. [3]. Log odds ratio was used to approximate a linear relationship between time (from stroke onset to treatment) and effect, and this gives a time-effect relationship shown below:

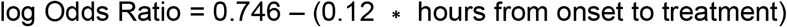

Maximum theoretical probability of good outcome (if treatment given at time of stroke onset) is 64.4%, as calculated by extrapolation of treatment effect back to t = 0 after stroke onset. By applying this decay, the benefit reduces to no clinical effect at 6.3 hours.

The probability of an additional good outcome given IVT is calculated. Good outcomes due to the baseline rate of good outcomes are not included.

Pseudocode:

~~~
# Set time where all effect of IVT is lost (6.3 hrs)
IVT_time_no_effect = 6.3 * 60
# Set maximum permitted time for IVT (4.5 hrs)
maximum_permitted_time_to_IVT = 4.5 * 60
# Set limits of max/min probability of good outcome
p_good_nLVO_IVT_max = 0.6444 # at t=0 after stroke onset
p_good_nLVO_IVT_min = p_good_nLVO_untreated
# Convert probabilities of good outcome to odds ratio
odds_good_nLVO_IVT_max =
   p_good_nLVO_IVT_max / (1 - p_good_nLVO_IVT_max)
odds_good_nLVO_IVT_min =
   p_good_nLVO_IVT_min / (1 - p_good_nLVO_IVT_min)
# Calculate fraction of time to no-effect at time of treatment
fraction_max_IVT_treatment_time_used =
   onset_to_needle / IVT_time_no_effect
# Calculate odds of good outcome based on time to treatment
max_effect_log_odds =
   log(odds_good_nLVO_IVT_max) - log(odds_good_nLVO_IVT_min)
odds_good_nLVO_IVT =
   exp(log(odds_good_nLVO_IVT_max) -
   (max_effect_log_odds * fraction_max_IVT_treatment_time_used))
# Convert odds of good outcome to probability of good outcome
p_good_nLVO_IVT = odds_good_nLVO_IVT / (1 + odds_good_nLVO_IVT)
# Set to min good outcome probability if time > max permitted time
if onset_to_needle > maximum_permitted_time_to_IVT:
   p_good_nLVO_IVT = p_good_nLVO_IVT_min
# Ensure all patients have at least minimum of good probability
if p_good_nLVO_IVT < p_good_nLVO_IVT_min:
   p_good_nLVO_IVT = p_good_nLVO_IVT_min
# Calculate probability of additional good outcome due to IVT
p_add_good_nLVO_IVT = p_good_nLVO_IVT - p_good_nLVO_IVT_min
~~~

#### 2.4.2. Calculate additional good outcomes for LVO (for those eligible for treatment)

Additional good outcomes for LVO may be the result of IVT or MT,

##### 2.4.2.1. Calculate additional good outcomes for LVO given IVT

As with the treatment of nLVO with IVT, the maximum effect and decay of effect of IVT (alteplase) was taken from the meta-analysis by Emberson *et al*.[3]. Log odds ratio was used to approximate a linear relationship between time (from stroke onset to treatment) and effect, and this gives a time-effect relationship shown below:

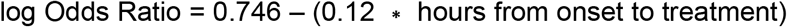

Maximum theoretical probability of good outcome (if treatment given at time of stroke onset) is 24.4%, as calculated by extrapolation of treatment effect back to t = 0 after stroke onset. By applying this decay, tbenefit reduces to no clinical effect at 6.3 hours.

The probability of an additional good outcome given IVT is calculated. Good outcomes due to the baseline rate of good outcomes are not included.

Pseudocode:

~~~
# Set time where all effect of IVT is lost (6.3 hrs)
IVT_time_no_effect = 6.3 * 60
# Set maximum permitted time for IVT (4.5 hrs)
maximum_permitted_time_to_IVT = 4.5 * 60
# Set limits of max/min probability of good outcome
p_good_LVO_IVT_max = 0.2441 # at t=0 after stroke onset
p_good_LVO_IVT_min = p_good_nLVO_untreated
# Convert probabilities of good outcome to odds ratio
odds_good_LVO_IVT_max =
   p_good_LVO_IVT_max / (1 - p_good_LVO_IVT_max)
odds_good_LVO_IVT_min =
   p_good_LVO_IVT_min / (1 - p_good_LVO_IVT_min)
# Calculate fraction of time to no-effect at time of treatment
fraction_max_IVT_treatment_time_used =
   onset_to_needle / IVT_time_no_effect
# Calculate odds of good outcome based on time to treatment
max_effect_log_odds =
   log(odds_good_LVO_IVT_max) - log(odds_good_LVO_IVT_min)
odds_good_LVO_IVT =
   exp(log(odds_good_LVO_IVT_max) -
   (max_effect_log_odds * fraction_max_IVT_treatment_time_used))
# Convert odds of good outcome to probability of good outcome
p_good_LVO_IVT = odds_good_LVO_IVT / (1 + odds_good_LVO_IVT)
# Set to min good outcome probability if time > max permitted time
if onset_to_needle > maximum_permitted_time_to_IVT:
   p_good_LVO_IVT = p_good_LVO_IVT_min
# Ensure all patients have at least minimum of good probability
if p_good_LVO_IVT < p_good_LVO_IVT_min:
   p_good_LVO_IVT = p_good_LVO_IVT_min
# Calculate probability of additional good outcome due to IVT
p_add_good_LVO_IVT = p_good_LVO_IVT - p_good_LVO_IVT_min
~~~

##### 2.4.2.2. Calculate additional good outcomes for LVO given MT

Those patients with LVO who do not respond to IVT (or who are unsuitable for IVT) may progress to MT.

The decay of effect of MT was taken from the Mr CLEAN multi-unit trial analysis <u>[19]</u>. This does not include patients who may be selected at later times with advanced imaging techniques. Log odds ratio was used to approximate a linear relationship between time (from stroke onset to treatment) and effect, and this gives a time-effect relationship as shown below:

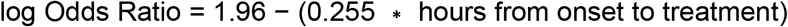

Maximum theoretical probability of good outcome, if treatment given at time of stroke onset, is 52.1 %, as calculated by extrapolation of treatment effect back to t = 0 after stroke onset. Applying this decay, the benefit reduces to no clinical effect at 8 hours.

The probability of an additional good outcome given MT is calculated. Good outcomes due to the baseline rate of good outcomes are not included.

Pseudocode:

~~~
# Set time where all effect of MT is lost
MT_time_no_effect = 8 * 60
# Set maximum permitted time for MT (6 hrs)
maximum_permitted_time_to_MT = 6 * 60
# Set limits of max and min probability of good outcome
p_good_LVO_MT_max = 0.5208 # at t=0 after stroke onset
p_good_LVO_MT_min = p_good_nLVO_untreated
# Convert probabilities of good outcome to odds ratios
odds_good_LVO_MT_max =
   odds_good_LVO_MT_max / (1 - odds_good_LVO_MT_max)
odds_good_LVO_MT_min =
   odds_good_LVO_MT_min / (1 - odds_good_LVO_MT_min)
# Calculate fraction of time to no-effect at time of treatment
fraction_max_MT_treatment_time_used =
   onset_to_puncture / MT_time_no_effect
# Calculate odds of good outcome based on time to treatment
max_effect_log_odds =
   log(odds_good_LVO_MT_max) - log(odds_good_LVO_MT_min)
odds_good_LVO_MT =
   exp(log(odds_good_LVO_MT_max) -
   (max_effect_log_odds * fraction_max_MT_treatment_time_used))
# Convert odds of good outcome to probability of good outcomes
p_good_LVO_MT = odds_good_LVO_MT / (1 + odds_good_LVO_MT)
# Set to min good outcome probability if time > max permitted time
if onset_to_puncture > maximum_permitted_time_to_MT:
   p_good_LVO_MT = p_good_LVO_MT_min
# Ensure all patients have at least minimum of good probability
if p_good_LVO_MT < p_good_LVO_MT_min:
   p_good_LVO_MT = p_good_LVO_MT_min
# Calculate probability of additional good outcome due to MT
p_add_good_LVO_MT = p_good_LVO_MT - p_good_LVO_MT_min
~~~

### 2.5. Calculate the weighted probability of good outcomes

Calculate the weighted good outcomes for each group my multiplying the probability of good outcomes by the fraction of patients in each group

~~~
# Good outcomes in stroke mimic patients
weighted_good_mimic = proportion_mimic * p_good_mimic
# Good outcomes in ICH
weighted_good_ICH = proportion_ICH * p_good_ICH
# Baseline good outcomes in nLVO
weighted_good_nLVO_base = proportion_nLVO * p_good_nLVO_untreated
# Additional outcomes for nLVO given IVT
weighted_good_nLVO_IVT =
   proportion_nLVO * nLVO_eligible_for_treatment *
   p_add_good_nLVO_IVT
# Baseline good outcomes in LVO
weighted_good_LVO_base = proportion_LVO * p_good_LVO_untreated
# Additional outcomes for LVO given IVT
weighted_good_LVO_IVT =
   proportion_LVO * LVO_eligible_for_treatment *
   p_add_good_LVO_IVT
~~~

For treatment of LVO with MT, the number of LVO patients treated with MT is reduced by the number of LVO patients who would have a good response to IVT (to avoid double-counting good outcomes for those patients receiving both IVT and MT).

~~~
# Additional outcomes for LVO given MT
prop_LVO_receiving_MT =
   (LVO_eligible_for_IVT * LVO_IVT_eligible_MT) –
   p_add_good_LVO_IVT)
weighted_good_LVO_MT =
   proportion_LVO * prop_LVO_receiving_MT *
   p_add_good_LVO_MT
~~~

### 2.6. Calculate overall probability of good outcomes

After calculation of baseline and treated outcomes, the overall proportion of good outcomes is calculated by summing the weighted proportion of good outcomes for each group.

Pseudocode:

~~~
overall_probability_good_outcomes =
   weighted_good_mimic +
   weighted_good_ICH +
   weighted_good_nLVO_base +
   weighted_good_nLVO_IVT +
   weighted_good_LVO_base +
   weighted_good_LVO_IVT +
   weighted_good_LVO_MT
~~~

### 2.7. Alternative MT effect decay

An alternative decay of effect of MT may be taken from the HERMES meta-analysis[20], using the common odds ratio of improved outcome. This includes patients who may be selected at later times with advanced imaging techniques, which may overestimate the overall effect of MT at longer onset-to-treatment times. Log odds ratio was used to approximate a linear relationship between time and effect, and this gives a time-effect relationship as shown below:

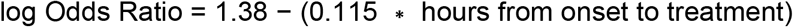

The maximum theoretical probability of good outcome (if treatment is given at time of stroke onset) is 37.8%, as calculated by extrapolation of treatment effect back to t = 0 after stroke onset. Applying this decay, the benefit reduces to no clinical benefit as 12 hours.

Note: though the theoretical t = 0 effect of Mr CLEAN vs. HERMES appears significantly different (52.1% vs 37.8%), the effect at realistic onset-to-treatment times are more similar, for example at 3 hours the predicted proportion of good outcomes for Mr CLEAN and HERMES are 33.6% and 30.1% respectively.

## 3. Results

Figure 2 shows the relationship between time to treatment for IVT and MT and outcomes, for eligible patients (those who receive IVT and/or MT). Without reperfusion treatments, the baseline number of good outcomes (mRS 0-1) is 321 / 1,000 admissions. With reperfusion, there is a theoretical maximum benefit of 219 additional good outcomes / 1,000 stroke patients with LVO. This would only occur if treatment were given immediately on stroke onset, and all patients were eligible to receive both IVT and MT. The equivalent theoretical maximum benefit for nLVO stroke patients is 125 additional good outcomes.

**Figure 2.**
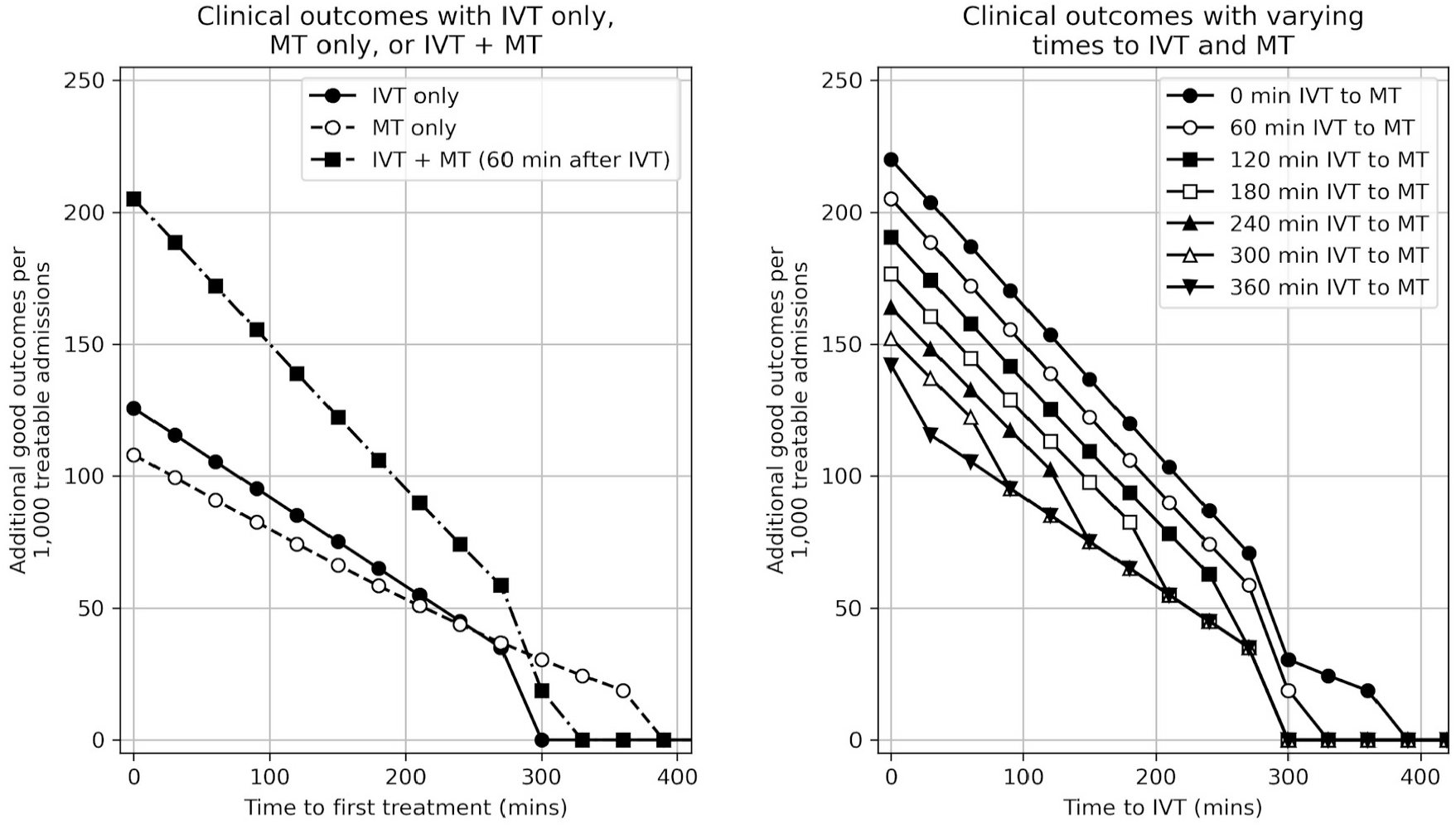
The relationship between time-to-treatment and outcome for treatable patients. Left panel: The number of additional good outcomes / 1,000 treatable patients if given IVT only, MT only, or IVT + MT (MT given only to LVO patients, with MT given 60 minutes after IVT). Right panel: The number of additional good outcomes / 1,000 treatable patients by time to IVT and time between IVT and MT (for LVO patients).

This theoretical maximum benefit is reduced by two factors. First, benefit declines as onset to treatment time increases - it is unrealistic to expect more than a small proportion of patients to receive treatment within 90 minutes of stroke onset. Second, not all patients are eligible for treatment. We adjust the benefit according to time to IVT and/or MT, and we restrict the maximum proportion of patients considered eligible for treatment. In all results presented here we have assumed maximum IVT and MT treatment rates of 20% and 10% respectively.

Applying these proportions of patients eligible for treatment yields a theoretical maximum benefit (if treatment were given immediately after stroke onset) of 57 additional good outcomes / 1,000 unselected stroke admissions. If IVT were to be given 60 minutes after stroke onset and MT were given (to LVO patients) 60 minutes later then the number of additional good outcomes would be 44 / 1,000 admissions.

## 4. Discussion

Here we provide calculations, pseudocode and a link to Python code for calculation of stroke outcomes based on times to thrombolysis and thrombectomy.

Some limitations of this approach should be noted:

- The effectiveness of IVT is modelled on alteplase. It is possible that other forms of recombinant tPA such as tenecteplase[21] could have different profiles.
- The data set reported for MT outcomes is currently smaller than that available for IVT, and so there is more uncertainty around the exact benefit of MT.
- Our method/code focuses on those patients who are able to receive MT within 8 hours. There is further benefit of MT to a smaller set of patients who do not fall into this time limit, but who have had advanced imaging demonstrating that a portion of the brain is still salvageable by MT[22–24].
- Outcome is provided as a dichotomised *good* vs. *not-good*. This misses beneficial effects where a patient may have an improved outcome, but not to the extent of reaching the threshold of *‘good’*. We use this dichotomised outcome as that was the design of the clinical trials.
- Outcome is not broken down into subgroups (e.g. by age cut off), and we assume all patients have the same time cut-off for allowing treatment (in some places, those aged 80 or more will not be treated with IVT after 3 hours have elapsed from onset of stroke, compared with 4.5 hours for those aged under 80).

Notwithstanding these limitations, modelling outcomes after IVT and/or MT, given the time to treatment, allows for different configuration of services to be modelled. For example, a *drip and ship* model will reduce average time to IVT but increase average time to MT, whereas a *mothership* model will reduce average time to MT but at the cost of average time to IVT.

## Data Availability

The code is available at https://github.com/MichaelAllen1966/stroke_outcome_algorithm

## Ethics

This paper provides a theoretical framework for estimation of clinical benefit from thrombolysis and thrombectomy, based only on published clinical trial data. No ethical approval was required.

## Competing interests

There are no competing interests to declare.

## Funding

This study was funded by the National Institute for Health Research (NIHR) Applied Research Collaboration (ARC) South West Peninsula. The views and opinions expressed in this paper are those of the authors, and not necessarily those of the NHS, the National Institute for Health Research, or the Department of Health.

## Data availability

The code is available at https://github.com/MichaelAllen1966/stroke_outcome_algorithm

## Notes

### Competing Interest Statement

The authors have declared no competing interest.

### Summary of Updates

Corrected section headings.

## References

1 Sentinel Stroke National Audit Programme - Annual Report 2019. HQIP. 2019. https://www.hqip.org.uk/resource/sentinel-stroke-national-audit-programme-annual-report-2019/ (accessed 13 Jul 2020).

2 Intercollegiate Stroke Working Party. National Clinical Guideline for Stroke, 5th Edition (Royal College of Physicians of London). 2016. www.strokeaudit.org.uk/guideline (accessed 2 Aug 2017).

3 Emberson J, Lees KR, Lyden P, et al. Effect of treatment delay, age, and stroke severity on the effects of intravenous thrombolysis with alteplase for acute ischaemic stroke: A meta-analysis of individual patient data from randomised trials. The Lancet 2014; 384: 1929–1935. doi:10.1016/S0140-6736(14)60584-5

4 Morris S, Hunter RM, Ramsay AIG, et al. Impact of centralising acute stroke services in English metropolitan areas on mortality and length of hospital stay: difference-in-differences analysis. Bmj 2014; 349: g4757–g4757. doi:10.1136/bmj.g4757

5 de la Ossa Herrero N, Carrera D, Gorchs M, et al. Design and Validation of a Prehospital Stroke Scale to Predict Large Arterial Occlusion The Rapid Arterial Occlusion Evaluation Scale. Stroke J Cereb Circ 2013; 45. doi:10.1161/STROKEAHA.113.003071

6 Flynn D, Francis R, Halvorsrud K, et al. Intra-arterial mechanical thrombectomy stent retrievers and aspiration devices in the treatment of acute ischaemic stroke : A systematic review and meta-analysis with trial sequential analysis. 2017; 2: 308–318. doi:10.1177/2396987317719362

7 Goyal M, Menon BK, Van Zwam WH, et al. Endovascular thrombectomy after large-vessel ischaemic stroke: A meta-analysis of individual patient data from five randomised trials. The Lancet 2016; 387: 1723–1731. doi:10.1016/S0140-6736(16)00163-X

8 Fransen PSS, Berkhemer OA, Lingsma HF, et al. Time to Reperfusion and Treatment Effect for Acute Ischemic Stroke: A Randomized Clinical Trial. JAMA Neurol 2016; 73: 190–6. doi:10.1001/jamaneurol.2015.3886

9 McMeekin P, White P, James MA, et al. Estimating the number of UK stroke patients eligible for endovascular thrombectomy. Eur Stroke J 2017; 2: 319–326. doi:10.1177/2396987317733343

10 White PM, Bhalla A, Dinsmore J, et al. Standards for providing safe acute ischaemic stroke thrombectomy services (September 2015). Clin Radiol 2017; 72: 175.e1–175.e9. doi:10.1016/j.crad.2016.11.008

11 Ismail M, Armoiry X, Tau N, et al. Mothership versus drip and ship for thrombectomy in patients who had an acute stroke: a systematic review and meta-analysis. J NeuroInterventional Surg 2019; 11: 14–9. doi:10.1136/neurintsurg-2018-014249

12 Ciccone A, Berge E, Fischer U. Systematic review of organizational models for intra-arterial treatment of acute ischemic stroke. Int J Stroke 2019; 14: 12–22. doi:10.1177/1747493018806157

13 Holodinsky JK, Williamson TS, Demchuk AM, et al. Drip-and-Ship vs. Mothership: Modelling Stroke Patient Transport for All Suspected Large Vessel Occlusion Patients. JAMA Neurol 2018; 75: 1477–86. doi:10.1001/jamaneurol.2018.2424

14 Holodinsky JK, Williamson TS, Kamal N, et al. Drip and ship versus direct to comprehensive stroke center. Stroke 2017; 48: 233–238. doi:10.1161/STROKEAHA.116.014306

15 Bonita R, Beaglehole R. Modification of Rankin Scale: Recovery of motor function after stroke. Stroke J Cereb Circ 1988; 19: 1497–500.

16 Gibson LM, Whiteley W. The differential diagnosis of suspected stroke: a systematic review. J R Coll Physicians Edinb 2013; 43: 114–8. doi:10.4997/JRCPE.2013.205

17 Dawson A, Cloud GC, Pereira AC, et al. Stroke mimic diagnoses presenting to a hyperacute stroke unit. Clin Med Lond Engl 2016; 16: 423–426. doi:10.7861/clinmedicine.16-5-423

18 McClelland G, Rodgers H, Flynn D, et al. The frequency, characteristics and aetiology of stroke mimic presentations: a narrative review. Eur J Emerg Med Off J Eur Soc Emerg Med 2019; 26: 2–8. doi:10.1097/MEJ.0000000000000550

19 Fransen PSS, Beumer D, Berkhemer O a, et al. MR CLEAN, a multicenter randomized clinical trial of endovascular treatment for acute ischemic stroke in the Netherlands: study protocol for a randomized controlled trial. Trials 2014; 15: 343. doi:10.1186/1745-6215-15-343

20 Muir KW, White P. HERMES: Messenger for stroke interventional treatment. Lancet 2016; 387: 1695–7. doi:10.1016/S0140-6736(16)00351-2

21 Gurbel PA, Hayes K, Bliden KP, et al. The platelet-related effects of tenecteplase versus alteplase versus reteplase. Blood Coagul Fibrinolysis 2005; 16: 1–7.

22 Albers GW, Marks MP, Kemp S, et al. Thrombectomy for Stroke at 6 to 16 Hours with Selection by Perfusion Imaging. N Engl J Med 2018; 378: 708–18. doi:10.1056/NEJMoa1713973

23 Campbell BCV, Mitchell PJ, Kleinig TJ, et al. Endovascular Therapy for Ischemic Stroke with Perfusion-Imaging Selection. N Engl J Med 2015; 372: 1009–1018. doi:10.1056/NEJMoa1414792

24 Kidwell CS, Jahan R, Gornbein J, et al. A Trial of Imaging Selection and Endovascular Treatment for Ischemic Stroke. N Engl J Med 2013; 368. doi:10.1056/NEJMoa1212793

